# Plasma proteomics of seizure-associated changes in epilepsy

**DOI:** 10.1101/2025.02.04.25321586

**Authors:** Sarah Akel, Saman Hosseini Ashtiani, Mayuresh Anant Sarangdhar, Markus Axelsson, Johan Zelano

## Abstract

Fluid biomarkers are emerging as crucial markers for diagnosis and disease monitoring in neurology. Epilepsy remains an exception despite seizures being known to result in metabolic changes, inflammation, and altered brain protein levels. Insight into how seizures affect the blood proteome is greatly needed.

This cross-sectional study used plasma samples from adults aged 18-50 with epilepsy recruited at five different neurology outpatient clinics in region Västra Götaland, Sweden to the Prospective Regional Epilepsy Database and Biobank for Individualized Clinical Treatment (PREDICT). Patients were categorized based on the presence of seizures; those with seizures ≤1 year (*n*=88, median number of seizures for patients who have experienced seizures in the last two months = 1) and those seizure-free (*n*=88) for >1 year.

Plasma proteomics was performed using four Olink Explore384 panels to analyze a total of 1447 proteins. Normalized Protein eXpression (NPX) values were compared between seizure and seizure-free participants. Two machine learning models, one linear and one non-linear, identified protein expression differences associated with seizure status through feature selection, resulting in 51 unique discriminative proteins. Twenty-three proteins were considered top differentially expressed after FDR adjustment (*p*≤0.05), including neurofilament light and several cytokines. Protein-protein interaction (PPI) analysis identified several clusters among the 51 discriminative proteins, with the largest clusters primarily associated with inflammatory processes. In addition, machine-learning independent gene set enrichment analysis identified 34 gene sets, mainly related to immune and inflammatory processes, that were also enriched in association with seizures.

In this study, persons with epilepsy and seizures had different plasma protein profiles compared to those seizure-free, primarily suggesting altered inflammatory/immune processes. The findings fit well with the growing evidence of inflammation as a key process in epilepsy. In addition to new therapeutic targets, immune processes need further exploration as candidate biomarkers for the monitoring of treatment responses in epilepsy.

## Introduction

Epilepsy affects over 50 million people worldwide, out of which one-third continue to have seizures despite antiseizure medications.^1^ Drug-resistant epilepsy is not only linked to seizure-related injury and death but also cognitive symptoms such as memory impairment.^2^

Blood protein levels are increasingly used in the study of brain disorders like Alzheimer’s disease and multiple sclerosis, particularly proteins linked to neurodegeneration and inflammation.^3–6^ Epileptic seizures leave short-term biochemical traces, such as lactic acidosis, but persistent biomarkers are not yet available.^7^ We have recently described that neurofilament-light (NfL) levels are higher if seizures persist in epilepsy.^8^ Emerging literature suggests that in addition to neuronal injury markers like NfL, cytokines increase following seizures.^9–11^ Based on this finding, along with evidence suggesting that drug-resistant epilepsy features chronic inflammation in affected brain tissue, we hypothesized that there are chronic alterations in the plasma proteome of patients with epilepsy with seizures compared to those who are seizure-free.^12,13^ Inflammatory mediators can, in turn, make neurons hyperexcitable, potentially creating a bidirectional interaction between blood and the brain in seizure-promoting states.^14,15^ This highlights the need for a more detailed understanding of changes in the plasma proteome induced by seizures. In addition to enhancing pathophysiological understanding, these insights could potentially offer scalable methods for monitoring disease activity and brain health in epilepsy through blood tests. Proteomic studies in epilepsy are emerging, but further research is still needed to address seizure-induced changes to the proteome.^16–19^

We analyzed 1447 plasma proteins in 176 individuals with epilepsy using four Olink Explore 384 panels. Our aim was to compare plasma protein profiles between persons with seizures and those who are seizure-free. Using machine learning, we identified 51 unique discriminative proteins between seizure and seizure-free groups. Among these, 23 proteins were considered the top differentially expressed, including the already known seizure-related NfL. Network and machine-learning independent enrichment analyses suggested that inflammatory processes were primarily associated with individuals who had recent seizures.

## Methods

### Study design and participant selection

**T**he Prospective Regional Epilepsy Database and Biobank for Individualized Clinical Treatment (PREDICT) (clinicaltrials.org, NCT04559919) aims to identify biomarkers of clinical utility in epilepsy and has recruited participants at five neurology outpatient clinics in Västra Götaland region of Sweden since December 2020. The cohort has a similar age and sex distribution to all adult patients seen at neurology departments for epilepsy in the region and Sweden as a whole.^19^ The study is approved by the Swedish Ethical Review Authority (approval number 2020-00853), and all participants provide written informed consent before inclusion. An overview of the study design is illustrated in Figure 1.

**Figure 1:**
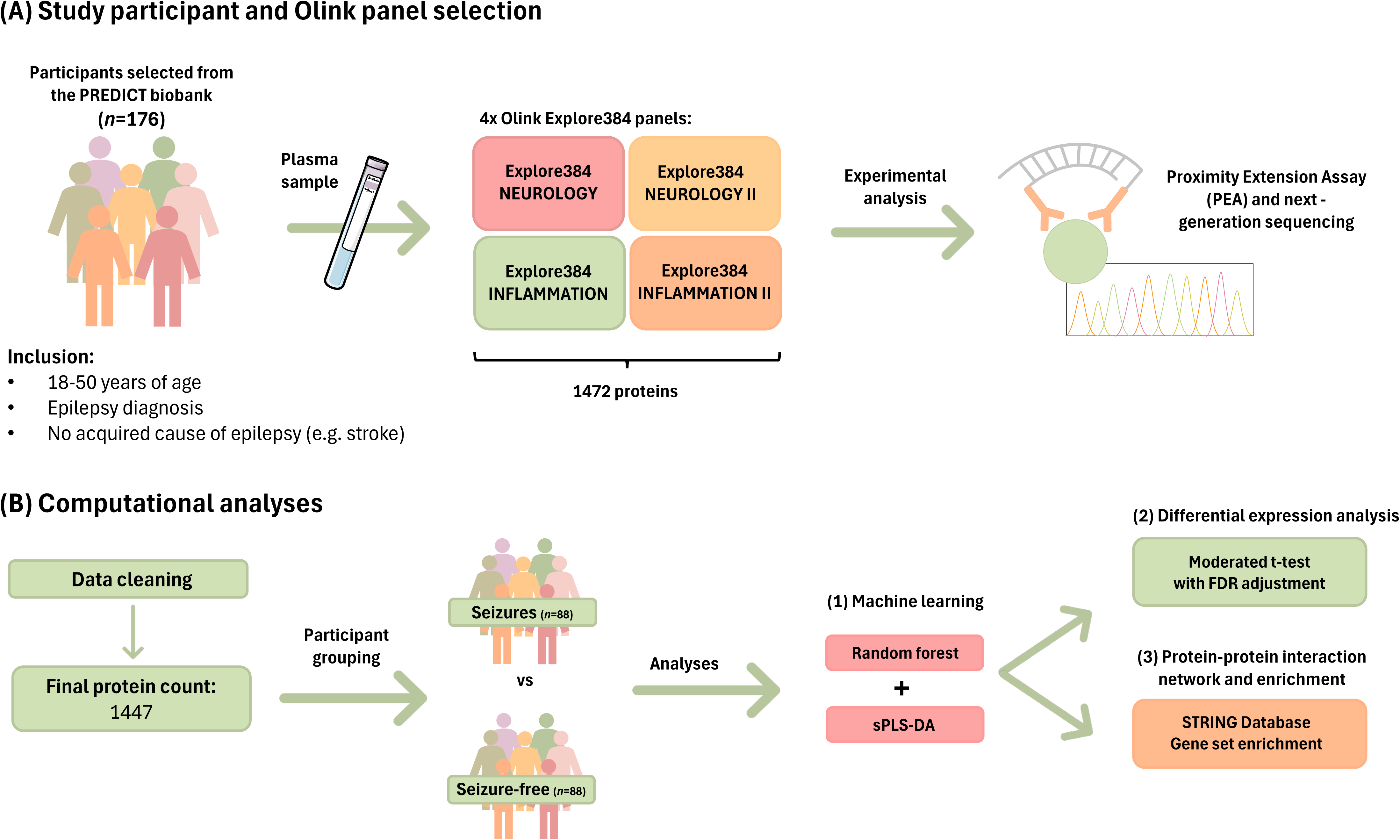
Overview of study design. (A) Plasma samples from 176 participants selected from the PREDICT biobank were analyzed using four Olink Explore384 panels. Experimental analysis was performed using the proximity extension assay in combination with next-generation sequencing. (B) A total of 1447 proteins underwent computational analysis where the values of these proteins were assessed in participants with seizures compared to participants who were seizure-free. Two machine learning models were utilized: Random forest and sPLS-DA. Further analysis involved a moderated t-test to find the top differentially expressed proteins selected from the machine learning models along with protein-protein interaction and enrichment analyses.

We included 176 participants from PREDICT, aged 18-50 years, with an epilepsy diagnosis as per the current ILAE definition^20^. The upper age limit was set to reduce variability from age-related brain changes. Participants were selected based on their seizure status. Participants who reported seizures ≤1 year at the last clinic visit/or blood sampling date (*n*=88) were categorized as “seizure” while participants who reported no seizures for >1 year (*n*=88) were categorized as “seizure-free”. All participants had their blood sampling within 10 days or less of the date of their seizure status information. None of the participants had an acquired cause of their epilepsy, such as stroke or TBI.

### Blood collection and storage

Upon recruitment to PREDICT, blood samples were collected into EDTA tubes and centrifuged for 10 minutes at 2000xg at room temperature. Plasma supernatant was collected and aliquoted, with all samples stored at -80°C in the regional healthcare-integrated Biobank Väst (registration number 890) until further analysis.

### Clinical data collection

Clinical data was collected into a pseudonymized clinical report form (CRF) from medical records and recruitment appointments by a neurologist. Variables used in this study include age, sex, radiology findings (clinical reports from MRI (*n*=130) and/or CT *n*=19)), seizure frequency and type of epilepsy (focal, generalized, or unknown onset).

### Olink Proteomics

Markers were quantified in plasma at SciLifeLab Uppsala (Uppsala University, Sweden) using four Olink Explore384 (Uppsala, Sweden) panels (Neurology, Neurology II, Inflammation, and Inflammation II). To fit the sample size within the capacity of two plates, a total of 176 participants were selected for this study. Olink uses a dual-recognition immunoassay based on their Proximity Extension Assay (PEA) method.^21^ Two oligonucleotide-coupled antibodies (PEA probes) bind closely to their protein target, enabling the oligonucleotides to hybridize. This creates a distinct DNA template quantified with next-generation sequencing. The output is expressed as Normalized Protein eXpression (NPX) values on a log2 scale indicating relative quantification.

### Data preprocessing

Data analyses were conducted in the R programming environment (version 4.4.1). We merged the four Olink panels, and in the case of overlapping assays (*n*=6) the assay with the highest Normalized Protein eXpression (NPX) value was kept in the resulting dataset. Nineteen proteins failed to meet Olink’s quality control criteria and were excluded. The final dataset comprised 1447 proteins.

### Machine learning

To capture both linear and non-linear associations between protein abundances and seizure status, two machine learning approaches were applied: Sparse Partial Least Squares Discriminant Analysis (sPLS-DA) using the R package mixOmics (version 6.28.0) ^22^ and the Random Forest R package (version 4.7-1.2).^23^

#### Sparse Partial Least Squares Discriminant Analysis (sPLS-DA)

sPLS-DA was employed to achieve both dimension reduction and feature selection, identifying proteins that best-differentiated participants with seizure vs. seizure-freedom. This method constructs components that maximize covariance between the response variable (seizure status) and predictors (protein levels). The optimal number of variables to keep was determined using grid search and repeated cross-validation. Through the mentioned model tuning, the balanced error rate (BER) criterion was used to choose the number of variables to select in the model (Figure S1A).

#### Random Forest

Complementary to the sPLS-DA model, we also employed Random Forest (RF). This ensemble method leverages multiple decision trees to enhance predictive accuracy and mitigate overfitting. Each tree was developed using a bootstrap sample, with a subset of features randomly chosen at each node to determine splits. The model was designed to classify participants based on seizure status, incorporating age and sex as predictors. Using 5,000 trees, we set the number of proteins considered at each split to 14, determined through a preliminary grid search using cross-validation for improving model performance (Figure S1B). We selected an equal number of proteins used by the RF model as the sPLS-DA model for classification to keep the balance in capturing linearity and non-linearity.

#### Model validation

Model performance metrics, including sensitivity, specificity, and area under the ROC curve, were calculated to assess classification performance for each model (Figure S1C). Feature importance was determined for the RF model using the mean decrease in impurity and for sPLS-DA using the loading weights of selected features.

### Statistical analysis

Using the moderated t-test from the limma R package (version 3.58.1^24,25^), we identified proteins selected from the union of sPLS-DA and RF that were differentially expressed between participant groups after FDR adjustment (adjusted *p* ≤ 0.05), controlling for age and sex. We also re-ran the same machine learning analysis and moderated t-test by categorizing participants into focal and generalized epilepsy subgroups to determine whether the proteomic profiles differed between these two types of onset.

### Protein-protein interaction network

A protein-protein interaction (PPI) network was generated using the STRING database (accessed 6^th^ December 2024, v12.0).^26^ The input gene set included the 51 proteins identified via the union of feature selection from sPLS-DA and RF models. We applied the default interaction confidence score threshold (0.4). We visualized the first shell of interactors in the network, consisting of up to 20 interactor proteins per query protein, to increase the network’s size and improve clustering and enrichment analyses. Markov Cluster Algorithm (MCL) was applied to identify protein clusters (inflation parameter = 4). Functional enrichment analyses were performed on the clusters (FDR adjusted *p*-value ≤ 0.05).

#### Sub-group analysis: focal and generalized epilepsy

A PPI network with clustering (MCL, inflation parameter = 1.5) was also constructed for proteins identified as classifiers of seizure status in both focal epilepsy and generalized. Functional enrichment analyses were performed on the clusters (FDR adjusted *p*-value ≤ 0.05). No additional first shell interactors were included.

### Gene set enrichment analysis (GSEA)

GSEA was conducted using GSEA v4.3.3 (Broad Institute, Cambridge, MA, USA).^27^ The NPX values of 1447 proteins were provided as input in a gct file. Patient phenotype information, categorized as ‘with seizure’ and ‘seizure-free,’ was supplied in a cls file. P-values for the enrichment scores were calculated through an empirical, phenotype-based permutation test. The number of permutations was set to 1,000, and the permutation seed was set to 149. Weighted enrichment statistics were used to calculate the enrichment scores. The C5 ontology gene set database was used as input which consists of 16,107 gene sets, and all other parameters were kept at their default settings. Gene sets with nominal *p*-value <0.05 and FDR *q*-value <25% were considered statistically significant enriched sets.

## Results

### Cohort description

The median age was 31 (range: 18-50) of the 176 participants, and most participants in the seizure group had focal epilepsy (Table 1). The majority of participants had normal radiology (*n*=104) or a non-acquired abnormality unrelated to the epilepsy (*n*=24) on imaging. There were 17 participants with an epileptogenic lesion on imaging. Thirty individuals had no clinical radiology reports, most of whom had generalized epilepsy (*n*=18).

**Table 1:**
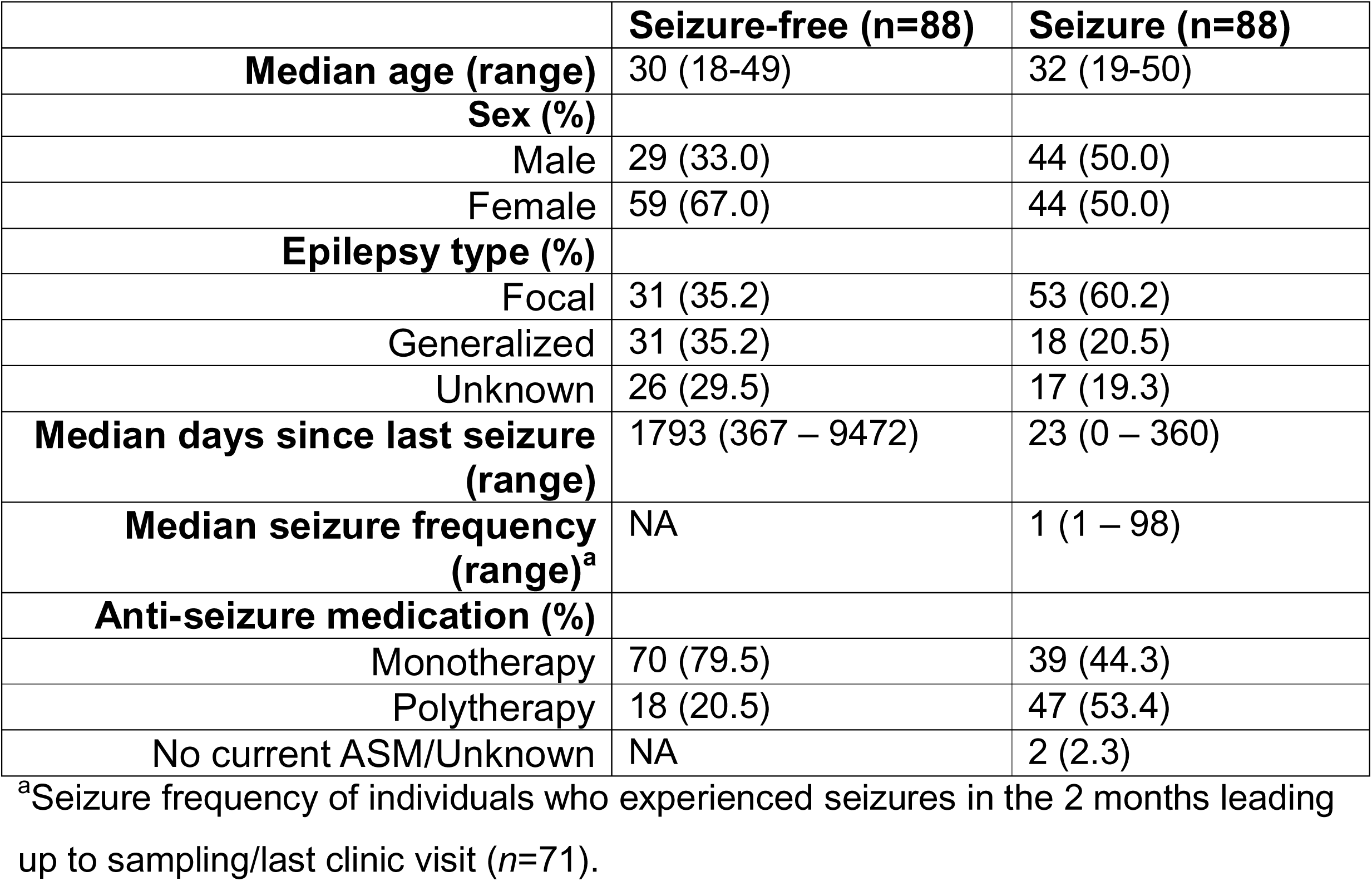
Participant demographics and characteristics (*n*=176)

|  | Seizure-free (n=88) | Seizure (n=88) |
| --- | --- | --- |
| **Median age (range)** | 30 (18-49) | 32 (19-50) |
| **Sex (%)** |  |  |
| Male | 29 (33.0) | 44 (50.0) |
| Female | 59 (67.0) | 44 (50.0) |
| **Epilepsy type (%)** |  |  |
| Focal | 31 (35.2) | 53 (60.2) |
| Generalized | 31 (35.2) | 18 (20.5) |
| Unknown | 26 (29.5) | 17 (19.3) |
| **Median days since last seizure (range)** | 1793 (367 – 9472) | 23 (0 – 360) |
| **Median seizure frequency (range)<sup>a</sup>** | NA | 1 (1 – 98) |
| **Anti-seizure medication (%)** |  |  |
| Monotherapy | 70 (79.5) | 39 (44.3) |
| Polytherapy | 18 (20.5) | 47 (53.4) |
| No current ASM/Unknown | NA | 2 (2.3) |

<sup>a</sup>Seizure frequency of individuals who experienced seizures in the 2 months leading up to sampling/last clinic visit ( $n=71$ ).

### Machine learning identifies proteins associated with seizures

Out of the 1447 proteins, sPLS-DA and RF each identified 30 proteins as important classifiers. Fifty-one of the 60 proteins were unique (Table S1), and 23 were considered top differentially expressed after a moderated t-test with FDR adjustment (Table 2, Figure 2). Samples are projected onto the two-dimensional space by their first two components calculated using sPLS-DA (Figure S1D).

**Figure 2:**
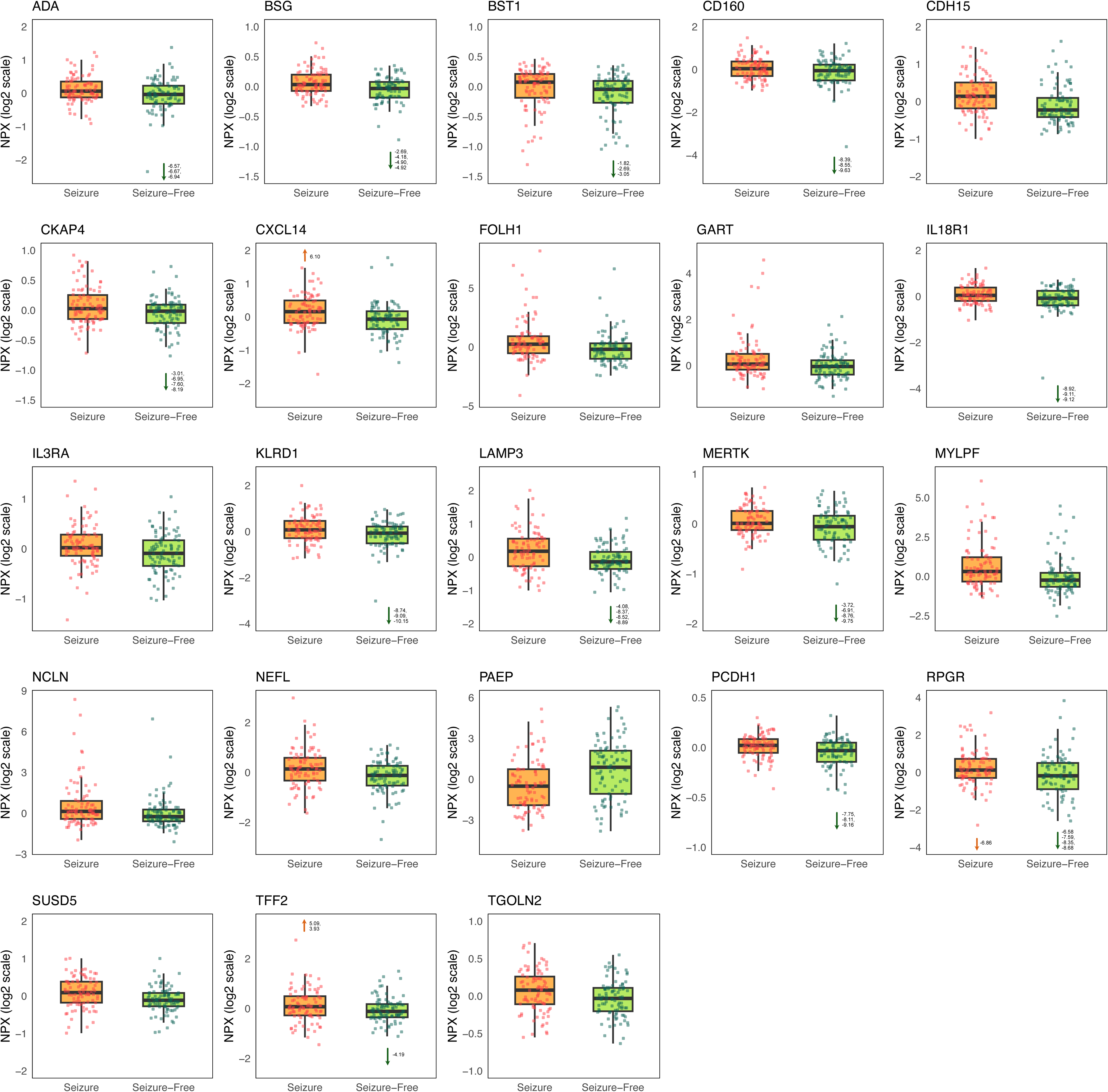
Tukey box and whisker plot illustrating the top 23 significantly different proteins between seizure and seizure-free groups. The median is represented by the center line of the box and whisker plot. The lower and upper hinges represent the 25th and 75th percentiles, respectively, while the whiskers extend 1.5 times the interquartile range from both hinges. Analysis was done using a moderated t-test with FDR adjustment (p≤0.05) and performed on the 51 discriminative proteins identified through machine learning analysis. Arrows depict the direction of values that appear outside of the visualized plot range.

**Table 2:** Description of top differentially expressed proteins identified via moderated t-test with FDR adjustment.

| Uniprot Accession | Gene symbol | Protein name | Log fold change | p-value<sup>a</sup> |
| --- | --- | --- | --- | --- |
| **Q9UQV4** | LAMP3 | Lysosome-associated membrane glycoprotein 3 | 0,636 | 0,001 |
| **Q96A32** | MYLPF | Myosin regulatory light chain 2, skeletal muscle isoform | 0,634 | 0,001 |
| **P35613** | BSG | Basigin | 0,335 | 0,002 |
| **Q07065** | CKAP4 | Cytoskeleton-associated protein 4 | 0,487 | 0,002 |
| **O95971** | CD160 | CD160 antigen | 0,577 | 0,003 |
| **P22102** | GART | Trifunctional purine biosynthetic protein adenosine-3 | 0,373 | 0,003 |
| **Q13241** | KLRD1 | Natural killer cells antigen CD94 | 0,591 | 0,003 |
| **P00813** | ADA | Adenosine deaminase | 0,429 | 0,004 |
| **P55291** | CDH15 | Cadherin-15 | 0,197 | 0,005 |
| **Q10588** | BST1 | ADP-ribosyl cyclase/cyclic ADP-ribose hydrolase 2 | 0,202 | 0,006 |
| **Q13478** | IL18R1 | Interleukin-18 receptor 1 | 0,534 | 0,006 |
| **Q03403** | TFF2 | Trefoil factor 2 | 0,346 | 0,006 |
| **Q04609** | FOLH1 | Glutamate carboxypeptidase 2 | 0,687 | 0,007 |
| **Q12866** | MERTK | Tyrosine-protein kinase Mer | 0,487 | 0,007 |
| **P07196** | NfL | Neurofilament light polypeptide | 0,299 | 0,007 |
| **P09466** | PAEP | Glycodelin | -0,694 | 0,007 |
| **Q92834** | RPGR | X-linked retinitis pigmentosa GTPase regulator | 0,676 | 0,007 |
| **O60279** | SUSD5 | Sushi domain-containing protein 5 | 0,154 | 0,009 |
| **Q969V3** | NCLN<sup>b</sup> | Nicalin | 0,592 | 0,01 |
| **O43493** | TGOLN2 | Trans-Golgi network integral membrane protein 2 | 0,11 | 0,011 |
| **O95715** | CXCL14<sup>b</sup> | C-X-C motif chemokine 14 | 0,249 | 0,013 |
| **Q08174** | PCDH1 | Protocadherin-1 | 0,402 | 0,015 |
| **P26951** | IL3RA | Interleukin-3 receptor subunit alpha | 0,143 | 0,022 |

<sup>a</sup>These p-values remained significant after FDR adjustment ( $p \leq 0.05$ ).
<sup>b</sup>Proteins with >30% of samples below the limit of detection (see Table S8).

### Protein-protein interaction network and enrichment analysis

The PPI network of the 51 unique proteins identified by sPLS-DA and RF comprised 71 nodes and 291 edges (Figure 3A). We identified ten clusters within the network (Table S4). The three largest clusters were primarily involved in inflammatory processes. Cluster 1, comprising 18 proteins, was enriched for “T cell activation” and “cytokine-cytokine receptor interaction”. Cluster 2 (13 proteins) was enriched for the “regulation of natural killer cell-mediated cytotoxicity”, “antigen processing and presentation” as well as “MHC protein complex binding” while cluster 3 (10 proteins) was enriched for “regulation of macrophage activation” and the regulation of cytokine production (including IL-1, IL-6, and tumor necrosis factor). Several small clusters were generated; cluster 4 was enriched for metabolism, while cluster 5 was enriched for cell adhesion. The remaining clusters had limited or no significant enrichment.

**Figure 3:**
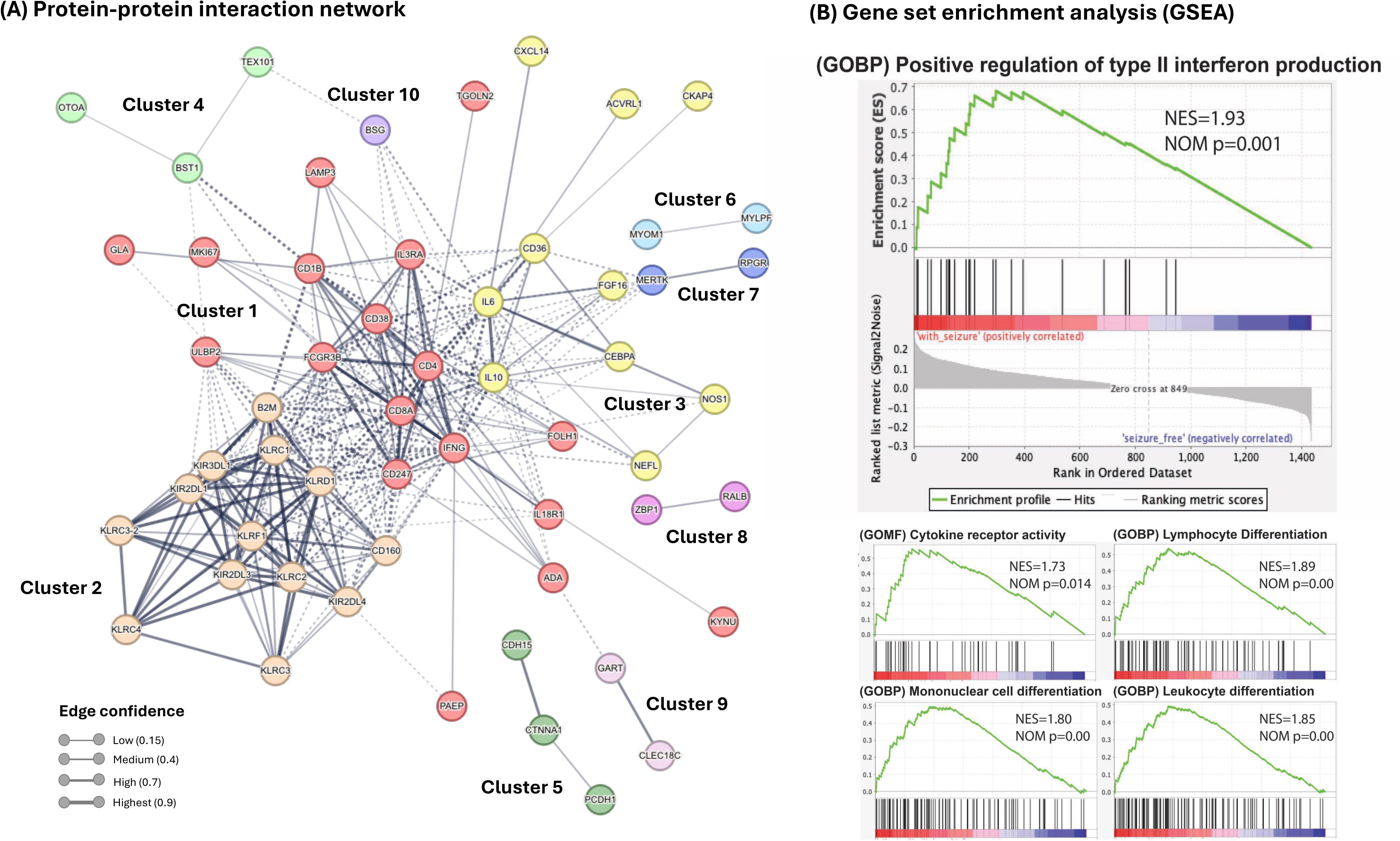
Protein-protein network and gene set enrichment analysis. (A) Protein-protein interaction network with MCL clustering of proteins determined as discriminative from machine learning models. Only proteins with connected nodes were visualized, and first shell interactors (up to 20 interactors per query protein) were included. Nodes represent proteins, and edges represent protein-protein interactions. Dashed lines indicate edges between clusters. (B) Representative gene set enrichment analysis (GSEA) plots. GSEA was based on protein profiles of “with seizure” (in red) and “seizure-free” (in blue) participants. NES: Normalized Enrichment Score, NOM p: nominal p-value

### Gene Set Enrichment Analysis (GSEA)

Complementary to the machine learning approach, we used GSEA software to identify gene sets associated with seizures. GSEA analysis revealed altered pathways related to seizures, with a total of 34 gene sets showing significant enrichment. These gene sets predominantly fell under immune (particularly T cell differentiation) and inflammation categories (such as type II interferon production)(Figure 3B, Table S5).

### Focal and generalized epilepsy

We also performed subgroup analyses on focal (*n*=84) and generalized (*n*=49) epilepsy. The two machine learning models identified 92 proteins as classifiers of seizure status in focal epilepsy, and 160 in generalized. Moderated t-test identified 26 proteins significantly different between seizure and seizure-free groups with focal epilepsy (adjusted *p*-value ≤0.05) (Table S2), while no proteins remained significant after FDR adjustment for generalized epilepsy (Table S3). Thirteen proteins overlapped between focal and generalized epilepsy, 23 between focal and all participants (focal, generalized + unknown onset), and 20 between generalized and all. A PPI network with clustering analysis was generated separately for focal and generalized epilepsy. Both networks showed one dominant cluster (focal = 42 proteins, generalized = 98 proteins) enriched for various inflammatory processes (Focal: Figure S2, Table S6; Generalized: Figure S3, Table S7). There were several shared inflammatory processes between focal and generalized protein networks such as “regulation of cytokine production” and “T cell activation”.

## Discussion

### Key findings

We used four Olink proteomic panels covering 1447 to study plasma changes associated with seizures in persons with epilepsy. Machine learning identified several discriminative proteins, of which 23 proteins were considered the top differentially expressed after FDR adjustment. Our findings demonstrate that the plasma proteome differs in persons with seizures compared to those with seizure-freedom, expanding the potential for future biomarkers beyond known short-term blood alterations after seizures, like lactic acidosis and muscle enzyme release. The findings are in agreement with several more recent reports of changes in neurological or inflammatory markers after seizures.^28^ Our proteomic approach identified several key markers, including the well-reported NfL, as well as various inflammatory markers such as interleukins (IL3RA, IL18R1) and other established inflammation markers (e.g., KLRD1 and KLRF1).

### PPI and enrichment analyses

Enrichment and protein-protein interaction analyses of the machine-learning identified proteins pointed to several inflammatory processes. The central three clusters were predominantly associated with inflammatory processes, including T cell activation, regulation of cytokine production, and natural killer cell-mediated toxicity. The protein network also identified IL-6 and IFNy as neighbor proteins interacting with our core protein network. Several studies have reported that they increase in epilepsy, with IL-6 in particular having been associated with seizure severity.^29–33^ As a confirmatory analysis, a machine learning-independent GSEA also demonstrated that immune and inflammation-related pathways were overrepresented in seizure cases compared to seizure-free cases. The analysis pointed to both adaptive and innate immune responses.

The interaction between brain seizure activity and inflammation is gaining interest not only in immune-mediated epilepsy or encephalitis, but also with the notion of inflammation perpetuating and maintaining an ictogenic state.^14^ Inflammation is also of interest in drug-resistant epilepsy^12,34^ Clinical trials of anti-inflammatory treatments in drug-resistant epilepsy have shown variable results, but a substantial proportion do respond.^35–37^ A more detailed understanding of the inflammatory pathways involved through proteomics could perhaps assist in predicting responses and selecting the right immunotherapy for appropriate patients.

Our method also identified NfL as a top differentially expressed protein, which has been previously reported by several investigators to increase after seizures.^9,11^ Traditionally recognized as a marker of axonal injury, NfL has been well-studied in conditions such as Alzheimer’s disease, amyotrophic lateral sclerosis (ALS), and multiple sclerosis (MS).^38–41^ However, higher levels may also indicate other processes, such as plasticity, rather than solely injury.^42,43^ In this study, we also found three other proteins associated with seizures as in our previous study: CDH15, PAEP, and PHOSPHO1.^19^

We also analyzed focal and generalized epilepsy separately, although the power of the analysis naturally decreased with participant numbers. The exact protein lists identified by machine-learning differed, but several GO terms related to inflammatory processes were common to both groups. One possible interpretation is that inflammation detected in the proteome indicates reactions to epileptic brain activity, rather than underlying epileptogenic processes, which are different in the two types of epilepsy. Larger studies are needed to explore this observation further, but if confirmed, it suggests that some inflammatory mediators may be seizure biomarkers in both focal and generalized epilepsy.

### Strengths and limitations

We used a dual machine-learning approach for complementary strengths of the respective algorithms. To capture complex dependency and interplay patterns between seizure status and protein NPX levels, we selected RF over gradient boosting methods, such as XGBoost, for its robustness to noise and overfitting and superior feature importance interpretability. RF’s bagging-based approach ensures stable and biologically interpretable feature selection in high-dimensional proteomic datasets. ^44^ Furthermore, empirical evidence supports the reliability of RF in biomarker discovery studies. ^44,45^ However, RF has inherent limitations concerning the reproducibility of feature importance scoring. The randomized nature of the algorithm, including the use of bootstrapped samples and random feature selection at each tree split, can result in variability in the feature importance scores between different runs. This variability may impact the consistency of the identified biomarkers, particularly in high-dimensional datasets with correlated features like proteomics data. To mitigate this limitation, we introduced a fixed random seed before each run of the RF model. Additionally, all analyses were performed on the same machine specifications to eliminate variability arising from differences in computational environments.

sPLS-DA was employed as a complementary method to RF to leverage its capacity for simultaneous feature selection and dimension reduction. By focusing on linear relationships, sPLS-DA provides an interpretable model that identifies a minimal set of proteins associated with seizure status while mitigating overfitting through sparsity. However, sPLS-DA’s limitations, such as its sensitivity to parameter tuning, assumption of linearity, and potential exclusion of moderate-effect features, should be noted. These characteristics underscore the importance of combining sPLS-DA with nonlinear approaches like RF to ensure a comprehensive understanding of the proteomic signatures associated with seizures.

Other limitations include the cohort heterogeneity. Although this was a large proteomic investigation in epilepsy, it is possible that the number of patients was too small for adequate statistical power to detect all significantly regulated proteins in persons with seizures. That a protein was not identified, should not be interpreted as having no difference in its expression caused by seizures; our aim was to perform a proof-of-concept analysis of the plasma proteome in persons with seizures. We used inclusion criteria focused on younger age and non-acquired epilepsy to reduce biological variability, but future proteomic studies are needed on specific epilepsies and patient groups. Reassuringly, some of the differentially expressed proteins have been identified in previous analyses.^8,19^ Among these are findings from a methodologically distinct mass spectrometry experiment, where we also found more inflammation in the plasma of patients with many seizures compared to those seizure-free.^46^ Another limitation is the use of Olink panels as a basis for the analysis – which focused on neurology and inflammation-related proteins. These were selected based on the pre-existing knowledge on inflammation in drug-resistant epilepsy, but naturally, additional proteomic studies that utilize larger panels containing a more biologically diverse set of proteins would be useful.

### Conclusion

We found differences in the plasma proteome between persons with epilepsy and seizures and those with epilepsy who are seizure-free. In addition to known markers like NfL, both machine-learning and machine-learning independent analyses identified inflammation-associated clusters. Longitudinal sampling would be an interesting next step, studying the dynamics of protein levels with different seizure frequencies in response to ASMs. Overall, our results suggest that there are protein alterations in the plasma of individuals with seizures, and these proteins may serve as potential markers in epilepsy.

## Data Availability

Results of the study contain sensitive personal data (according to Swedish research regulations)and cannot be shared by the authors.

## Acknowledgments

This study was conducted using professional biobank services from Biobank West and Biobank Sweden. We would like to thank the Affinity Proteomics unit (SciLifeLab) at Uppsala (Sweden) for the experimental analysis of the Olink panels. Figure 1 contains an icon provided by Servier Medical Art by Servier (https://smart.servier.com/), licensed under CC BY 4.0 (https://creativecommons.org/licenses/by/4.0/).

This study was funded by the Wallenberg Center for Molecular and Translational Medicine, the Jeansson foundation (JS2014-0032), Swedish Society for Medical Research (SS18-0040), Region Västra Götaland (VGFOUREG-968476), the ALF-agreement (ALFGBG-715781), and the Wilhelm & Martina Lundgrens Vetenskapsfond, Hjärnfonden, Amlöv foundation and Ann-Louise och Sven-Erik Beiglers foundation.

## References

1. Chen Z, Brodie MJ, Liew D, Kwan P. Treatment Outcomes in Patients With Newly Diagnosed Epilepsy Treated With Established and New Antiepileptic Drugs: A 30-Year Longitudinal Cohort Study. JAMA Neurol. Mar 1 2018;75(3):279–286. doi:10.1001/jamaneurol.2017.3949

2. Gourmaud S, Shou H, Irwin DJ, et al. Alzheimer-like amyloid and tau alterations associated with cognitive deficit in temporal lobe epilepsy. Brain : a journal of neurology. Jan 1 2020;143(1):191–209. doi:10.1093/brain/awz381

3. Pereira JB, Janelidze S, Smith R, et al. Plasma GFAP is an early marker of amyloid-beta but not tau pathology in Alzheimer’s disease. Brain : a journal of neurology. Dec 16 2021;144(11):3505–3516. doi:10.1093/brain/awab223

4. Mielke MM, Hagen CE, Wennberg AMV, et al. Association of Plasma Total Tau Level With Cognitive Decline and Risk of Mild Cognitive Impairment or Dementia in the Mayo Clinic Study on Aging. JAMA Neurol. Sep 1 2017;74(9):1073–1080. doi:10.1001/jamaneurol.2017.1359

5. Tiedt S, Duering M, Barro C, et al. Serum neurofilament light: A biomarker of neuroaxonal injury after ischemic stroke. Neurology. Oct 2 2018;91(14):e1338–e1347. doi:10.1212/WNL.0000000000006282

6. Kuhle J, Nourbakhsh B, Grant D, et al. Serum neurofilament is associated with progression of brain atrophy and disability in early MS. Neurology. Feb 28 2017;88(9):826–831. doi:10.1212/WNL.0000000000003653

7. Matz O, Zdebik C, Zechbauer S, et al. Lactate as a diagnostic marker in transient loss of consciousness. Seizure. Aug 2016;40:71–5. doi:10.1016/j.seizure.2016.06.014

8. Akel S, Asztely F, Banote RK, Axelsson M, Zetterberg H, Zelano J. Neurofilament light, glial fibrillary acidic protein, and tau in a regional epilepsy cohort: High plasma levels are rare but related to seizures. Epilepsia. 2023;10.1111/epi.17713

9. Giovannini G, Bedin R, Ferraro D, Vaudano AE, Mandrioli J, Meletti S. Serum neurofilament light as biomarker of seizure-related neuronal injury in status epilepticus. Epilepsia. Jan 2022;63(1):e23–e29. doi:10.1111/epi.17132

10. Nass RD, Akgün K, Elger C, et al. Serum biomarkers of cerebral cellular stress after self-limiting tonic clonic seizures: An exploratory study. Seizure. Feb 2021;85:1–5. doi:10.1016/j.seizure.2020.12.009

11. Akel S, Asztely F, Banote RK, Axelsson M, Zetterberg H, Zelano J. Neurofilament light, glial fibrillary acidic protein, and tau in a regional epilepsy cohort: High plasma levels are rare but related to seizures. Epilepsia. Oct 2023;64(10):2690–2700. doi:10.1111/epi.17713

12. Campos-Bedolla P, Feria-Romero I, Orozco-Suárez S. Factors not considered in the study of drug-resistant epilepsy: Drug-resistant epilepsy: Assessment of neuroinflammation. Epilepsia Open. Aug 2022;7 Suppl 1(Suppl 1):S68–s80. doi:10.1002/epi4.12590

13. Li J, Shi D, Wang L, Wu G. Chronic neuroinflammation regulates cAMP response element-binding protein in the formation of drug-resistant epilepsy by activating glial cells. Journal of Neurorestoratology. 2022/06/01/ 2022;10(2):100006. 10.1016/j.jnrt.2022.100006

14. Librizzi L, Vila Verde D, Colciaghi F, et al. Peripheral blood mononuclear cell activation sustains seizure activity. Epilepsia. Jul 2021;62(7):1715–1728. doi:10.1111/epi.16935

15. Hanin A, Cespedes J, Dorgham K, et al. Cytokines in New-Onset Refractory Status Epilepticus Predict Outcomes. Ann Neurol. Jul 2023;94(1):75–90. doi:10.1002/ana.26627

16. Pires G, Leitner D, Drummond E, et al. Proteomic differences in the hippocampus and cortex of epilepsy brain tissue. Brain Commun. 2021;3(2):fcab021. doi:10.1093/braincomms/fcab021

17. Sun J, Jiang T, Gu F, Ma D, Liang J. TMT-Based Proteomic Analysis of Plasma from Children with Rolandic Epilepsy. Dis Markers. 2020;2020:8840482. doi:10.1155/2020/8840482

18. Banote RK, Larsson D, Berger E, Kumlien E, Zelano J. Quantitative proteomic analysis to identify differentially expressed proteins in patients with epilepsy. Epilepsy Res. Aug 2021;174:106674. doi:10.1016/j.eplepsyres.2021.106674

19. Akel S, Banote RK, Asztely F, Zelano J. Protein profiling in plasma for biomarkers of seizure. Epilepsy Research. 2023/11/01/ 2023;197:107241. 10.1016/j.eplepsyres.2023.107241

20. Fisher RS, Acevedo C, Arzimanoglou A, et al. ILAE Official Report: A practical clinical definition of epilepsy. Epilepsia. 2014;55(4):475–482. 10.1111/epi.12550

21. Wik L, Nordberg N, Broberg J, et al. Proximity Extension Assay in Combination with Next-Generation Sequencing for High-throughput Proteome-wide Analysis. Mol Cell Proteomics. 2021;20:100168. doi:10.1016/j.mcpro.2021.100168

22. Rohart F, Gautier B, Singh A, Lê Cao KA. mixOmics: An R package for ‘omics feature selection and multiple data integration. PLOS Computational Biology. 2017/11// 2017;13(11):e1005752-e1005752. doi:10.1371/JOURNAL.PCBI.1005752

23. Liaw A, Wiener M. Classification and Regression by randomForest. R News. 2002;2:18-22. 3.

24. Phipson B, Lee S, Majewski IJ, et al. Robust hyperparameter estimation protects against hypervariable genes and improves power to detect differential expression. The Annals of Applied Statistics. 2016/06;10(2)doi:10.1214/16-AOAS920

25. Ritchie ME, Phipson B, Wu D, et al. limma powers differential expression analyses for RNA-sequencing and microarray studies. Nucleic Acids Research. 2015/4// 2015;43(7):e47-e47. doi:10.1093/NAR/GKV007

26. Szklarczyk D, Gable AL, Nastou KC, et al. The STRING database in 2021: customizable protein-protein networks, and functional characterization of user-uploaded gene/measurement sets. Nucleic Acids Res. Jan 8 2021;49(D1):D605–d612. doi:10.1093/nar/gkaa1074

27. Subramanian A, Tamayo P, Mootha VK, et al. Gene set enrichment analysis: a knowledge-based approach for interpreting genome-wide expression profiles. Proc Natl Acad Sci U S A. Oct 25 2005;102(43):15545–50. doi:10.1073/pnas.0506580102

28. Banote RK, Akel S, Zelano J. Blood biomarkers in epilepsy. Acta Neurol Scand. Oct 2022;146(4):362–368. doi:10.1111/ane.13616

29. Uludag IF, Duksal T, Tiftikcioglu BI, Zorlu Y, Ozkaya F, Kirkali G. IL-1β, IL-6 and IL1Ra levels in temporal lobe epilepsy. Seizure. Mar 2015;26:22–5. doi:10.1016/j.seizure.2015.01.009

30. Gao F, Gao Y, Zhang SJ, et al. Alteration of plasma cytokines in patients with active epilepsy. Acta Neurol Scand. Jun 2017;135(6):663–669. doi:10.1111/ane.12665

31. Ishikawa N, Kobayashi Y, Fujii Y, Kobayashi M. Increased interleukin-6 and high-sensitivity C-reactive protein levels in pediatric epilepsy patients with frequent, refractory generalized motor seizures. Seizure. Feb 2015;25:136–40. doi:10.1016/j.seizure.2014.10.007

32. Wang Y, Wang D, Guo D. Interictal cytokine levels were correlated to seizure severity of epileptic patients: a retrospective study on 1218 epileptic patients. J Transl Med. Dec 1 2015;13:378. doi:10.1186/s12967-015-0742-3

33. Mao L-Y, Ding J, Peng W-F, et al. Interictal interleukin-17A levels are elevated and correlate with seizure severity of epilepsy patients. Epilepsia. 2013/09/01 2013;54(9):e142–e145. 10.1111/epi.12337

34. Toledo A, Orozco-Suárez S, Rosetti M, et al. Temporal lobe epilepsy: Evaluation of central and systemic immune-inflammatory features associated with drug resistance. Seizure - European Journal of Epilepsy. 2021;91:447–455. doi:10.1016/j.seizure.2021.07.028

35. Falsaperla R, Collotta AD, Marino SD, et al. Drug resistant epilepsies: A multicentre case series of steroid therapy. Seizure. Feb 13 2024;117:115–125. doi:10.1016/j.seizure.2024.02.007

36. Kimizu T, Takahashi Y, Oboshi T, et al. Methylprednisolone pulse therapy in 31 patients with refractory epilepsy: A single-center retrospective analysis. Epilepsy Behav. Aug 2020;109:107116. doi:10.1016/j.yebeh.2020.107116

37. von Rhein B, Wagner J, Widman G, Malter MP, Elger CE, Helmstaedter C. Suspected antibody negative autoimmune limbic encephalitis: outcome of immunotherapy. Acta neurologica Scandinavica. Jan 2017;135(1):134–141. doi:10.1111/ane.12575

38. Jung Y, Damoiseaux JS. The potential of blood neurofilament light as a marker of neurodegeneration for Alzheimer’s disease. Brain. 2024;147(1):12–25. doi:10.1093/brain/awad267

39. Götze K, Vrillon A, Bouaziz-Amar E, et al. Plasma neurofilament light chain in memory clinic practice: Evidence from a real-life study. Neurobiology of Disease. 2023/01/01/ 2023;176:105937. 10.1016/j.nbd.2022.105937

40. Disanto G, Barro C, Benkert P, et al. Serum Neurofilament light: A biomarker of neuronal damage in multiple sclerosis. Annals of Neurology. 2017;81(6):857–870. 10.1002/ana.24954

41. Agah E, Mojtabavi H, Behkar A, et al. CSF and blood levels of Neurofilaments, T-Tau, P-Tau, and Abeta-42 in amyotrophic lateral sclerosis: a systematic review and meta-analysis. Journal of Translational Medicine. 2024/10/21 2024;22(1):953. doi:10.1186/s12967-024-05767-7

42. Sainio MT, Rasila T, Molchanova SM, et al. Neurofilament Light Regulates Axon Caliber, Synaptic Activity, and Organelle Trafficking in Cultured Human Motor Neurons. Front Cell Dev Biol. 2021;9:820105. doi:10.3389/fcell.2021.820105

43. Yuan A, Sershen H, Veeranna, et al. Neurofilament subunits are integral components of synapses and modulate neurotransmission and behavior in vivo. Mol Psychiatry. Aug 2015;20(8):986–94. doi:10.1038/mp.2015.45

44. Mohammed A, Kora R. A comprehensive review on ensemble deep learning: Opportunities and challenges. Journal of King Saud University - Computer and Information Sciences. 2023/02/01/ 2023;35(2):757–774. 10.1016/j.jksuci.2023.01.014

45. Acharjee A, Larkman J, Xu Y, Cardoso VR, Gkoutos GV. A random forest based biomarker discovery and power analysis framework for diagnostics research. BMC Medical Genomics. 2020/11/23 2020;13(1):178. doi:10.1186/s12920-020-00826-6

46. Ashtiani SH, Akel S, Berger E, Zelano J. Plasma proteomics in epilepsy: Network-based identification of proteins associated with seizures. Epilepsy Res. Nov 19 2024;209:107480. doi:10.1016/j.eplepsyres.2024.107480

