## Supplemental Figures for "Plasma proteomics of seizure-associated changes in epilepsy"

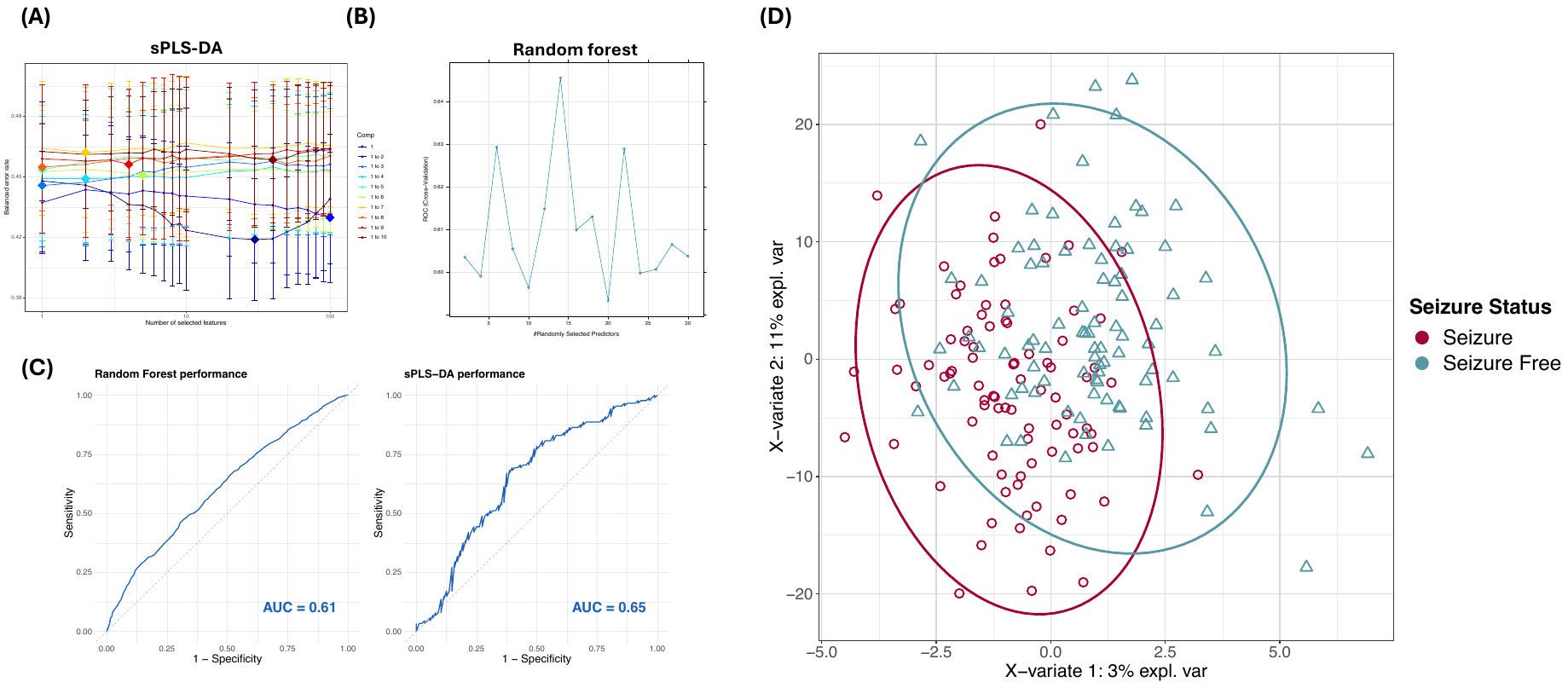

**Figure S1:** **Tuning and performance of machine-learning models.** (A) Balanced error rate (BER) criterion for choosing the number of selected variables in sPLS-DA, using cross validation and a grid of number of variables to go through. The BER increases with the addition of any number of dimensions, suggesting that one dimension is sufficient. The optimal number of variables is indicated with a diamond. (B) Using grid search and cross validation to find the optimal number of proteins at each split in the model. Y-axis indicates the average cross-validated Area Under the Curve (AUC) of the receiver operating characteristic (ROC) curves for different number of chosen proteins at each decision tree split. (C) ROC curves assessing classification accuracy of sPLS-DA and Random Forest models. (D) sPLS-DA plot based on the first two components reflecting the greatest variance. 95% confidence ellipses are depicted around individuals' groups.

**Figure S2: Protein-protein interaction network with MCL clustering for focal epilepsy**. Input proteins were the proteins determined as discriminative from machine learning models. Only proteins with connected nodes were visualized. Nodes represent proteins, and edges represent protein-protein interactions. Dashed lines indicate edges between clusters.

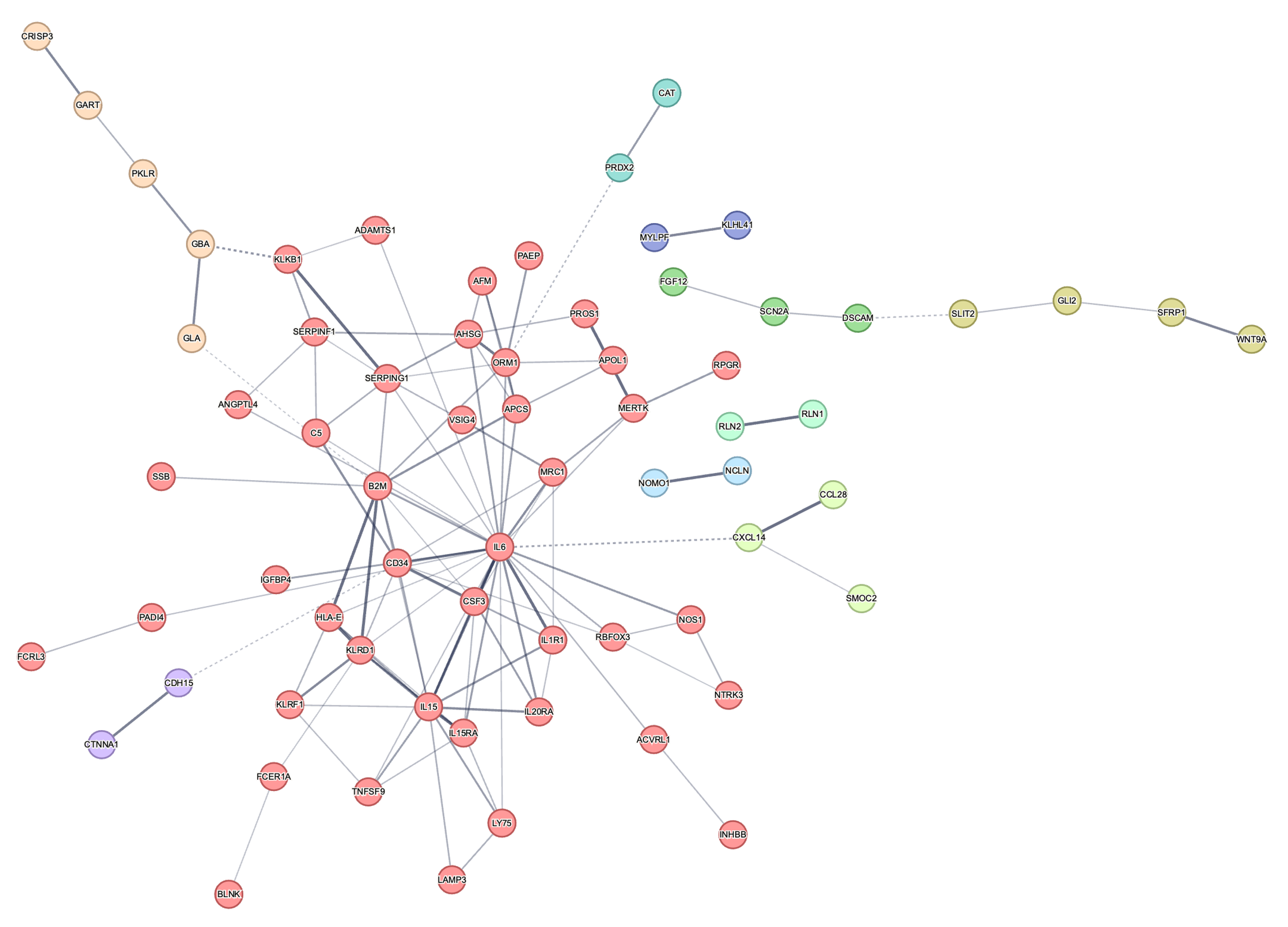

**Cluster 1**

**Cluster 2**

**Cluster 3**

**Cluster 4**

**Cluster 5**

**Cluster 6**

**Cluster 7**

**Cluster 8**

**Cluster 9**

**Cluster 10**

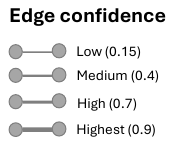

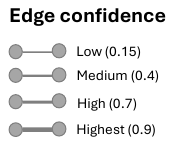

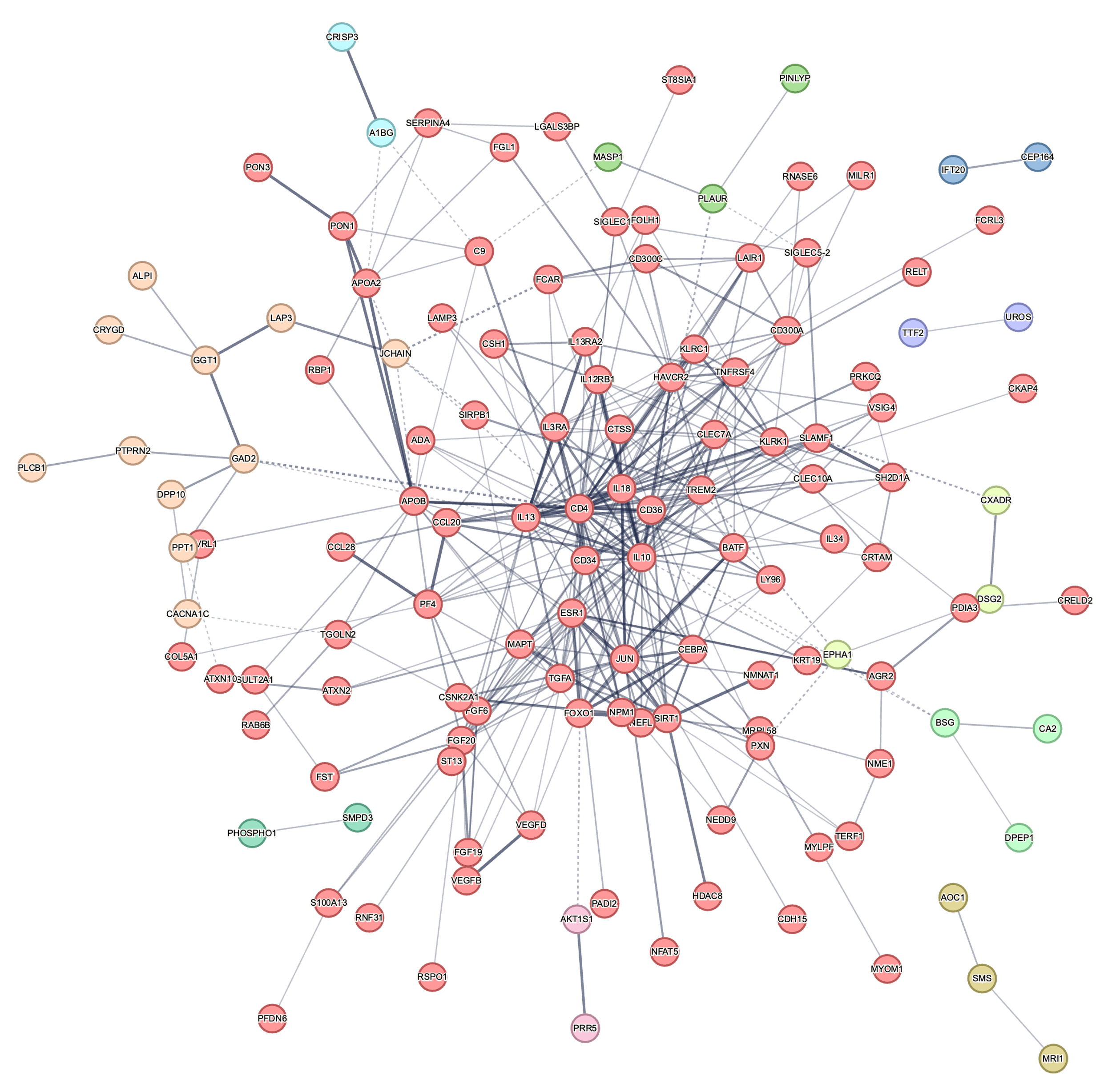

**Cluster 11**

**Cluster 10**

**Cluster 9**

**Cluster 8**

**Cluster 7**

**Cluster 6**

**Cluster 5**

**Cluster 1**

**Cluster 4**

**Cluster 3**

**Cluster 2**

**Figure S3: Protein-protein interaction network with MCL clustering for generalized epilepsy.** Input proteins were the proteins determined as discriminative from machine learning models. Only proteins with connected nodes were visualized. Nodes represent proteins, and edges represent protein-protein interactions. Dashed lines indicate edges between clusters.
